# Portable in-clinic video-based gait analysis: validation study on prosthetic users

**DOI:** 10.1101/2022.11.10.22282089

**Authors:** Anthony Cimorelli, Ankit Patel, Tasos Karakostas, R. James Cotton

## Abstract

Despite the common focus of gait in rehabilitation, there are few tools that allow quantitatively characterizing gait in the clinic. We recently described an algorithm, trained on a large dataset from our clinical gait analysis laboratory, which produces accurate cycle-by-cycle estimates of spatiotemporal gait parameters including step timing and walking velocity. Here, we demonstrate this system generalizes well to clinical care with a validation study on prosthetic users seen in therapy and outpatient clinic. Specifically, estimated walking velocity was similar to annotated 10-meter walking velocities, and cadence and foot contact times closely mirrored our wearable sensor measurements. Additionally, we found that a 2D keypoint detector pre-trained on largely able-bodied individuals struggles to localize prosthetic joints, particularly for those individuals with more proximal or bilateral amputations, but it is possible to train a prosthetic-specific joint detector. Further work is required to validate the other outputs from our algorithm including sagittal plane joint angles and step length. Code and trained weights will be released upon publication.

## Introduction

Gait impairments are a common target for rehabilitation. The most widely used outcome measures are the 10-meter walk test or 6-minute walk test which measure walking speed and endurance (1), but do not capture walking biomechanics. Alternatively, a motion analysis laboratory uses optical motion capture and force plates to obtain precise estimates of joint kinematics and kinetics and compute temporal and spatiotemporal gait parameters (2). However, the cost, time, and equipment required for formal gait analysis preclude performing it frequently, during clinical encounters, or outside the laboratory. There is a substantial need for clinically-usable tools that fill the gap between formal gait assessment and performance-based outcome measures to enable routine, quantitative characterization of gait and its associated impairments. These tools would enable more sensitive outcome measures to follow patients’ progress with therapy as well as to better power research to improve interventions. It would also enable routine screening of gait parameters in the clinic. This has many potential applications, such as allowing early detection of gait parameters associated with a risk of falling (3), which could then enable earlier interventions with fall prevention strategies.

Given the importance of analyzing gait and mobility, numerous approaches have been explored including wearable sensors and video analysis. Many different types of wearable sensors and algorithms have been described ranging from wrist devices that estimate step counts to placing numerous sensors over the body to estimate more complete kinematics (4–6). An advantage of wearables is they can enable ubiquitous monitoring of activity throughout the day (7). However, they can often require extensive and time-consuming calibration, require complicated and often proprietary algorithms to extract the relevant information, and have a wide range of reported accuracies (8).

In recent years, human pose estimation using deep learning has seen an extremely rapid advance yielding numerous approaches that can estimate 3D joint locations and pose (9– 11). However, the performance measure for the majority of this computer vision research is the accuracy of estimating 3D joint locations and not biomechanically motivated kinematics or gait parameters. Using multiple cameras, joint locations can be triangulated in 3D to produce more accurate estimates and these systems have been validated on numerous aspects of gait (12–18). In general, the need for multiple cameras makes these systems less portable and amenable for use in a clinic, although OpenCap has shown it is possible using only a computer and two smartphones with calibrated positions (19). Approaches have been developed that can analyze cycle-by-cycle gait from monocular video (20, 21), but they have not been validated on data acquired in the clinic or on clinical populations. Alternatively, other approaches train a neural network to directly map a sequence of 2D keypoints to average gait parameters that have been tested on clinical populations but do not enable analyzing individual gait cycles (22, 23).

We developed an algorithm, the Gait Transformer, trained on a large clinical gait laboratory dataset of paired videos and motion capture data (24). This Gait Transformer decomposes HPE trajectories of walking into individual gait cycles to produce accurate estimates of gait event timing and walking velocity, *when tested on that dataset*. However, artificial intelligence (AI) algorithms can be sensitive to changes in the data distribution. In this case, there are numerous differences between videos of gait collected in the real world or clinic and those from the gait laboratory. Thus, validating how this algorithm generalizes to data collected in clinical settings – the primary goal – is critical to enabling its use. The goals of this work include: 1) describe our combined system including smartphone application, wearable sensors, Pose Pipeline (25) and Gait Transformer (24) for clinical gait analysis, 2) validate the performance of this system on data acquired in the clinic, 3) identify under which conditions it performs well and when it is less reliable.

We performed this validation on prosthetic users and selected this population for several reasons. From a technical perspective, this is a challenging test of this system as the limbs of prosthetic users often appear visually different than able-bodied individuals, and it was previously unknown if pretrained HPE algorithms will generalize to prosthetic users. In addition, some prosthetic users walk with significant gait deviations compared to able-bodied individuals (26), which further challenges the Gait Transformer. From a clinical perspective, this is a population that would benefit from routine access to video-based gait analysis to monitor improvements in walking with therapy or with adjustments to prosthetic components. For example, routine, quantitative gait analysis could identify improvements in the quality of walking during therapy, such as increased symmetry and time spent in single stance on the prosthetic limb. This may be a more sensitive outcome measure, even if walking velocity has plateaued. This could enable physical therapists to demonstrate to insurance agencies that patients are still making progress and justify additional sessions. Or, more sensitive measures could provide data for prosthetists to show advanced components also improve quality of walking. In addition, these quantitative gait analysis could assist prosthetists with dynamic alignment during clinical visits.

## Methods

### Data Collection

#### Mobile Acquisition and Wearable Sensors

ideo and sensor data is acquired on an android smartphone using a custom app to synchronize recording from both modalities (**Fig 1**). The mobile phone is mounted on a 3-axis gimbal (DJI Osmo Mobile 2) to improve the video stability when following subjects during ambulation. The video was obtained in portrait orientation at 1080×1920 resolution at 30 frames per second.

**Fig. 1.**
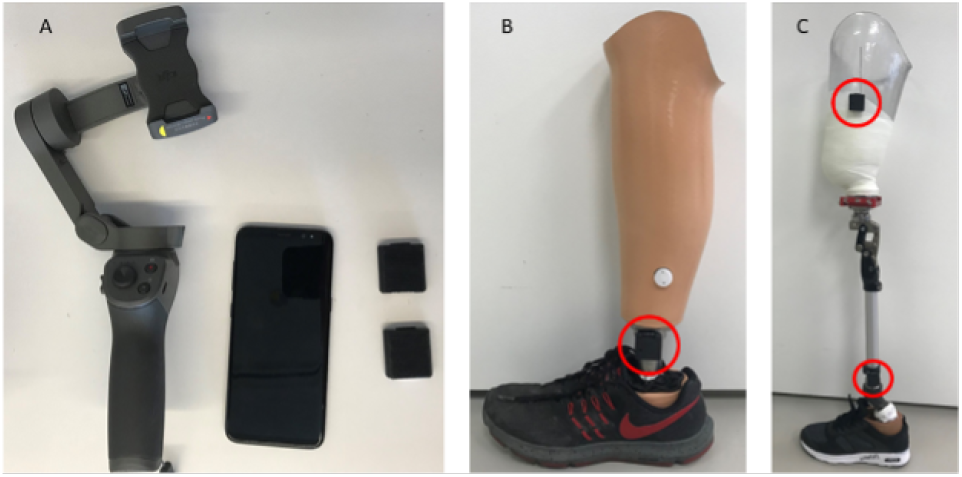
A) Android cell phone, two wearable sensors and gimbal. B) One sensor placed on the shank of a definitive transtibial prosthesis with adhesive Velcro. C) Two sensors placed on the shank/thigh of a diagnostic transfemoral.

The wearable sensors are a custom design and have previously been described (27, 28). They stream data from the IMU to the smartphone over Bluetooth Low-Energy (BLE). The IMU is an ICM-20948 and outputs 3-axis accelerometer and gyroscope data at 562.5 Hz. Magnetometer data is available but is not used in this study and we typically do not stream it in order to optimize the BLE bandwidth. The sensors can also acquire two channels of EMG data, but this was not used in these experiments. Prior to experiments, the magnetometer is calibrated in the location where data will be collected by rotating around each of the axes and accounting for hard and soft iron distortions. IMU data is fused on the sensor using a complementary filter to compute the 3D orientation, and the estimated orientation is streamed over BLE. Compared to our prior works using silicone-encapsulated wearable sensors attached to the skin, in this work the sensors were placed in a 3D-printed case with velcro on the outside for attachment to the body.

#### Clinical Population and Data Annotation

This study was approved by the Northwestern University Institutional Review Board. Video of gait and other activities was obtained from a convenience sample of lower-limb prosthetic users seen in an outpatient prosthetics clinic or participating in outpatient physical therapy at Shirley Ryan AbilityLab. For each subject, we recorded their age, height, level, and (bi)laterality of amputation, the etiology of the amputation, the type of prosthetic components, and their Medicare Functional Classification Level (K-level). Walking data was collected in either prosthetic or therapy clinics. In the case of therapy, videos were obtained as participants performed their usual therapy.

Sensors were placed on both the shank and thigh on the prosthetic limb(s), with up to four sensors used in the case of individuals with bilateral amputations. They were attached either with double-sided velcro attached to the prosthetic pylon, and socket (for transfemoral amputees) or with a velcro strap around the thigh. They were placed laterally, with the IMU Z axis pointed laterally and the X axis pointed down. In this work, only the data from the shank IMU was used to detect the prosthetic limb swinging.

Videos were annotated with the activity being performed (e.g., overground walking, treadmill walking, running, other therapeutic tasks), the view (frontal, sagittal, or a mixture), and the subjective accuracy of keypoint tracking of the prosthetic limb with a 3-point Likert scale ranging from 1 (poor) to 3 (good). Specifically 1 indicated the keypoints do not locate the prosthetic joints and tracking is frequently inaccurate, 2 indicated they locate the joint but are intermittently inaccurate, and 3 indicates the locate the joint well throughout the video. Whether the prosthetic was visible or occluded by clothing was also annotated for each video. Time points when the participant entered and exited 10-meter areas indicated by tape on the ground were also recorded to compute ground truth velocity.

#### Velocity Annotation

Tape was placed on the ground at 10-meter spacing in locations where subjects would typically walk such as the hallway in the prosthetic clinic and in multiple locations in the therapy gyms. The start and end times when subjects completed a straight overground walk between these markers were retrospectively annotated. This provided both ground truth measurements of their walking velocity and identified specific video segments where participants were walking, as the collected data contained a mixture of activities.

In this work, we focus our analysis on video segments where individuals are performing above-ground walking in a forward direction during the time window where they were walking between the two pieces of tape, which we refer to as timed walking segments. We focus on the 10-meter annotation periods both to determine the accuracy of the system when computing gait speed and because these were segments where individuals were known to be walking and were in view of the camera.

### Data Processing

#### Pose Pipe

The input to the gait transformer is a series of 3D keypoint locations. To obtain these, the video was processed with PosePipe (25), a human pose estimation pipeline based on DataJoint (29) that simplifies running cutting-edge HPE algorithms on video. The steps used in the pipeline include (1) a tracking algorithm (30) to compute bounding box tracks for all people in the scene followed by (2) manually annotating the bounding box for the subject of interest undergoing gait analysis. (3) Then we perform top-down 2D keypoints detection in each frame using the MMPose toolbox (31), specifically using an HRNet (32) trained on the COCO dataset (33) using distribution aware relative keypoints (34), (4) the 2D keypoint trajectories are then lifted to 3D joint locations (35).

We also used DeepLabCut (DLC) (36) to train a custom 2D keypoint detector for prosthetic users by manually annotating both intact and prosthetic hip, knee and ankle joints for a subset of videos where keypoint detection was performing poorly. We computed the 2D keypoints on those same videos with this model and replaced any prosthetic joints computed by MMPose with the estimates from DLC. This corrected set of keypoints was then passed to the lifting step and then to the gait transformer.

We flagged any frames as clipped whenever any of the key-points of the leg came within 10 pixels of the edge of the screen.

#### Gait Transformer

The sequence of 3D joint location was mapped onto the relative timing of four gait events (right and left foot contact and toe off) as well as the pelvis velocity using the Gait Transformer (24). This is trained on a large dataset of walking videos with synchronous marker-based motion capture and force plate data from our clinical gait laboratory. For training, data was aligned in the sagittal direction using the medial orientation of the pelvis. It also outputs sagittal plane joint kinematics including foot position relative to the pelvis, foot velocity, and hip and knee angles, which we do not focus on in this work. We refer the reader to the (24) for details of the architecture and training of the gait transformer. Compared to that work, we retrained the gait transformer and excluded bilateral elbows and wrists as we found occasionally the use of assistive devices (i.e., canes or crutches) in a less common pattern would trigger false detection of steps.

The Gait Transformer was applied to the lifted 3D joint trajectories. Training samples from the gait laboratory are typically only a few gait cycles long and we found that it did not generalize to inference on much longer sequences. Videos acquired in the clinic were much longer than a few gait cycles, so we applied it on a sliding window of 90 frames (3 seconds), which covers at least one complete stride for the majority of subjects. For each position of the sliding window, we used the middle output other than the beginning and end where we used the corresponding half of the sequence to pad the output.

#### Sensor processing

In order to validate the accuracy of the gait transformer event timing of the prosthetic limb, we used gyroscope data from the wearable sensor on the prosthetic shank to detect the prosthetic-limb swing phase. Sensor data is timestamped to the smartphone time. Gyroscope data were sampled at a nominal sampling rate of 562.5Hz. The Android system time of each Bluetooth packet is also stored and linear regression is used to calibrate the sensor timebase against the Android time, typically with an updated sampling rate of 1-2 Hz different than the nominal value.

We also noted that the video start timestamp showed some latency compared to the sensor timestamps. This has been resolved in more recent versions of our smartphone application with an API that acquires a more precise, per-frame timestamp. We used the hip and knee sagittal plane angles from the gait transformer to adjust for this timing error by finding the offset that minimized the mean squared error between the gyroscope on the shank and the change in that angle computed from the gait transformer outputs. This was typically around 170ms.

We detected a prosthetic limb swing from the gyroscope on the prosthetic shank. With sensors placed on the lateral side of the shank, the z-axis is roughly aligned to be perpendicular to the sagittal plane. We applied an 8th-order low-pass Chebyshev filter to the gyroscope with a cutoff of 35 Hz. All negative values were zeroed out and a median filter of 360ms was applied to deglitch a few strides where participants caught their toe and a brief reversal was seen in the gyroscope midtrace. These deglitched positive segments were identified as swings with the time the sign became negative identified as the start and end of swing periods.

#### Sensor versus video cadence

We compared the cadence estimated with the gait transformer over 10-meter walking segments to that computed from the sensors. We computed the cadence from video by averaging 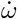 over the timed walking segment and converting from strides in rad/s to steps/min (i.e.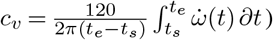). We computed cadence from the sensors using the average stride time of the prosthetic limb side over the steps in the time window: 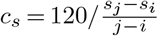, where *i* and *j* index the first and last sensor swing time, respective, that fall within the timed walking period.

#### Matching video and sensor foot contacts

We also compared the time when the end of swing from the gyroscopes to the foot contact time from the gait transformer. This analysis only reflects a bound, because detecting when the prosthetic shank stops rotating forward in swing (i.e., when the gyroscope swaps signs) approximates the end of swing time but is not the actual time the foot contacts the ground and will typically occur slightly before true foot contact (**Fig 2**). For each walking segment, we computed the offset that produced the closest matches between the end of prosthetic leg swings detected by the sensors and the time of prosthetic foot contact estimated from the Gait Transformer and Kalman filter (typically about 100ms). After this we measured the foot contact detection accuracy as the residual timing error between the offset sensor times and the detected video times. We also measured the fraction of events that were detected with a window of 500ms.

**Fig. 2.**
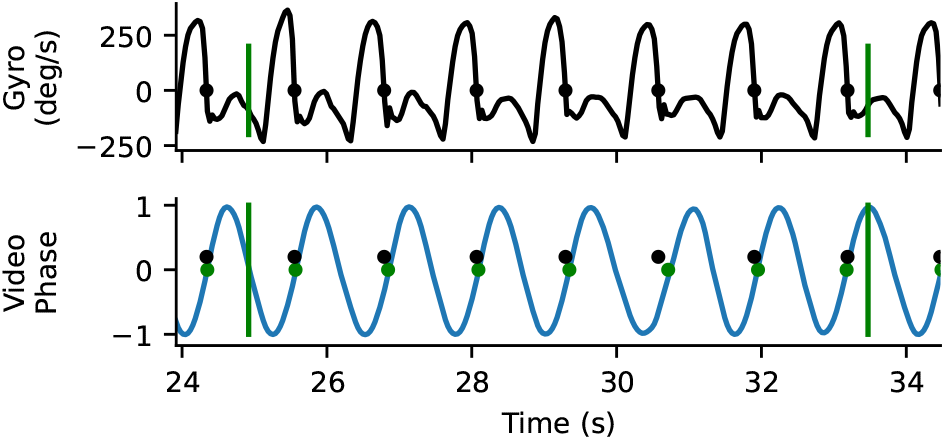
Example gyro and video timing information. Top trace is the z-component of the gyroscope mounted on the tibial shank. The time point where the gyroscope goes from positive to negative (where heel stops rotating forward relative to knee) is identified as a close proxy for foot contact. Bottom plot shows the sin component of the quadrature output for heel strike, with the positively directed zero crossing marker. There is a close correspondence between detected foot contacts from the gyro (blue) and the video (yellow). Vertical bars correspond to the annotated boundaries of the 10-meter walk test.

## Results

### Participant demographics

From 19 participants, we annotated 231 timed walking during level walking, with 79 in the frontal plane, 67 in the sagittal plane and the remainder either oblique or changing. When restricted to only the frontal plane, timed walking segments were obtained from a total of 16 participants.

**Table 1.**
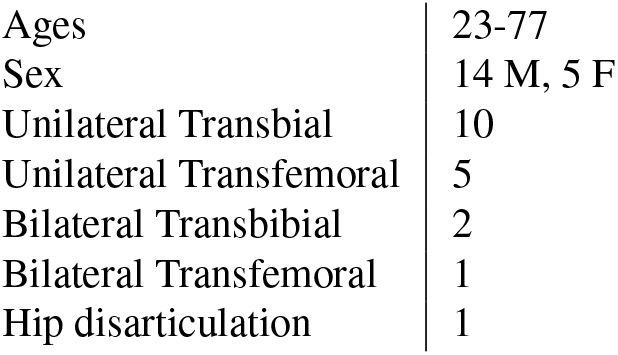
Demographic information of subjects

#### Usability of the system

Our system made it easy to collect gait data in clinical situations, including physical therapy and prosthetic appointments. Wearable sensors took less than 30 seconds to apply and remove and would require no setup time if not using sensors for validation studies. Data was easily and routinely obtained from subjects in both therapy and prosthetic appointments. This was particularly true for the frontal view, but obtaining clear sagittal views in hallways was often challenging due to space limitations. No subjects withdrew from the study. Example visualizations from the system are shown in Fig 3.

**Fig. 3.**
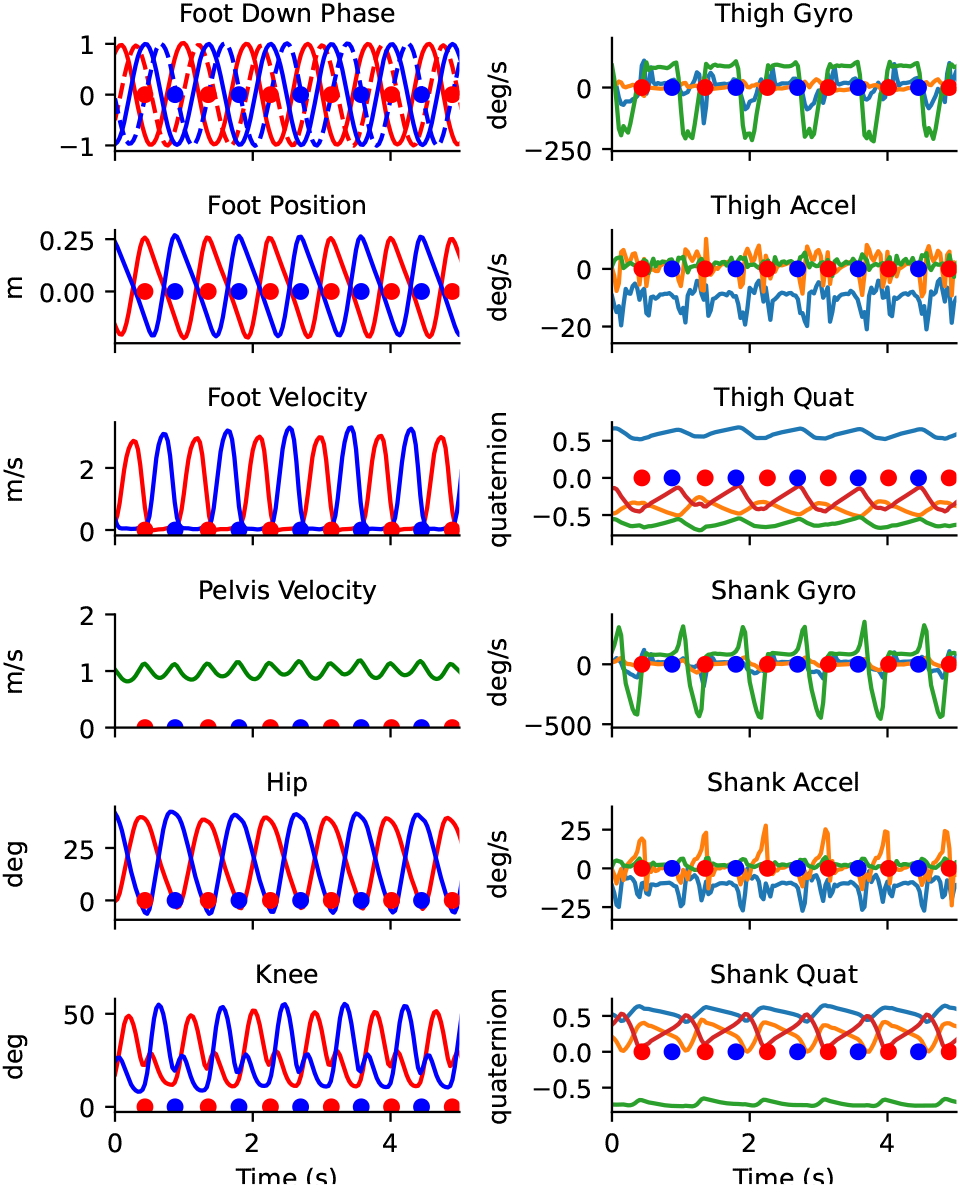
Traces from 5 seconds of walking from the portable system. The left column shows the outputs from the Gait Transformer and the right column shows the raw sensor data from the thigh and shank.

#### 2D Keypoint accuracy

Our 2D keypoint detection used an MMPose model, which is trained on the COCO dataset and primarily contains able-bodied individuals. We found that it frequently generalized to prosthetic users, but not always, and that quality of 2D keypoint tracking varied across subjects. In some cases, the detector would eschew localizing the prosthetic joint of the subject of interest and track the corresponding joint of a nearby therapist. While 2D keypoint detectors, including MMPose, output a per-joint confidence estimate, this has not been systematically tested for prosthetic users. For each video, we manually annotated our subjective estimate of tracking quality from 1 (poor) to 3 (good). For each timed walking segment, we also computed the average confidence that MMPose reported for the prosthetic ankle. **Fig 4a** shows the histogram of ankle keypoint qualities, which demonstrates that the confidence estimates from the keypoint detector align with our manual annotation. In fact, the interquartile ranges of ankle confidence conditioned on each of our annotated qualities were nearly non-overlapping (IQR for keypoint quality 1: [0.44,0.60]; 2: [0.59,0.80], 3: [0.77,0.87]).

**Fig. 4.**
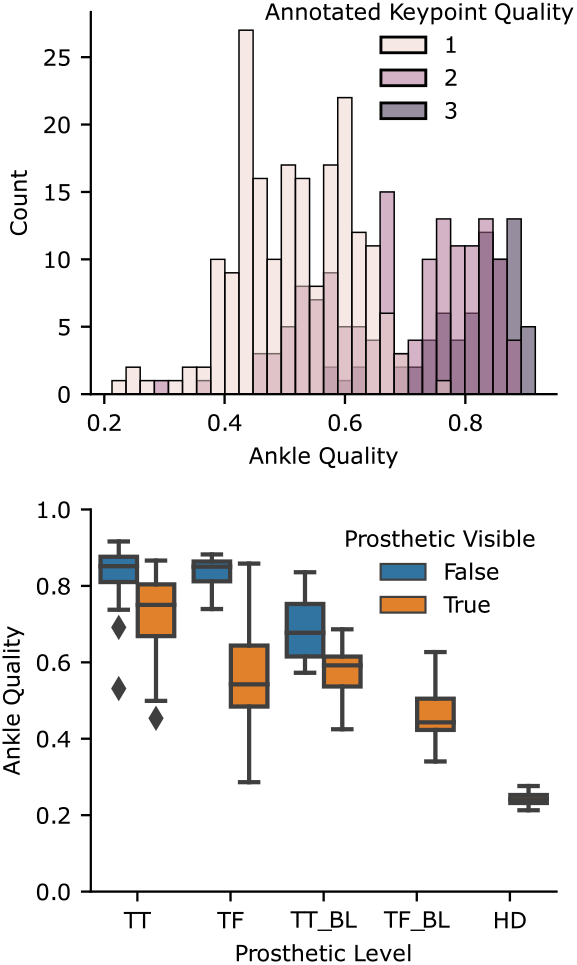
Accuracy of ankle keypoints for prosthetic users. The left panel shows the histogram of average ankle qualities for videos stratified by manual annotation of quality (with 3 being the best), showing that the estimated quality corresponded with our annotation. The right panel shows the average ankle quality statified by prosthetic level and whether the prosthetic was covered by clothing. TT: transtibial; TF: transfemoral; HD: hip-disarticulation; BL: bilateral

After validating that the ankle joint confidence is valid in prosthetic users, we then conditioned on the prosthetic level and whether clothing was covering the prosthesis or not, which revealed two trends (**Fig 4b**). Bilateral amputations or more proximal amputations resulted in worse tracking of the ankle joint. Ankle tracking was particularly poor in the case of a prosthetic user participant with a bilateral transfemoral amputation and another with a hip disarticulation.

In comparison to the MMPose model trained on COCO (33), our DLC model that was specifically trained on our prosthetic user was able to perform much better, **Fig 5**. Note that we did not test the generalization of this model to new users, and all prosthetic users we analyzed with our DLC model had 20 frames manually annotated and were included in the training dataset.

**Fig. 5.**
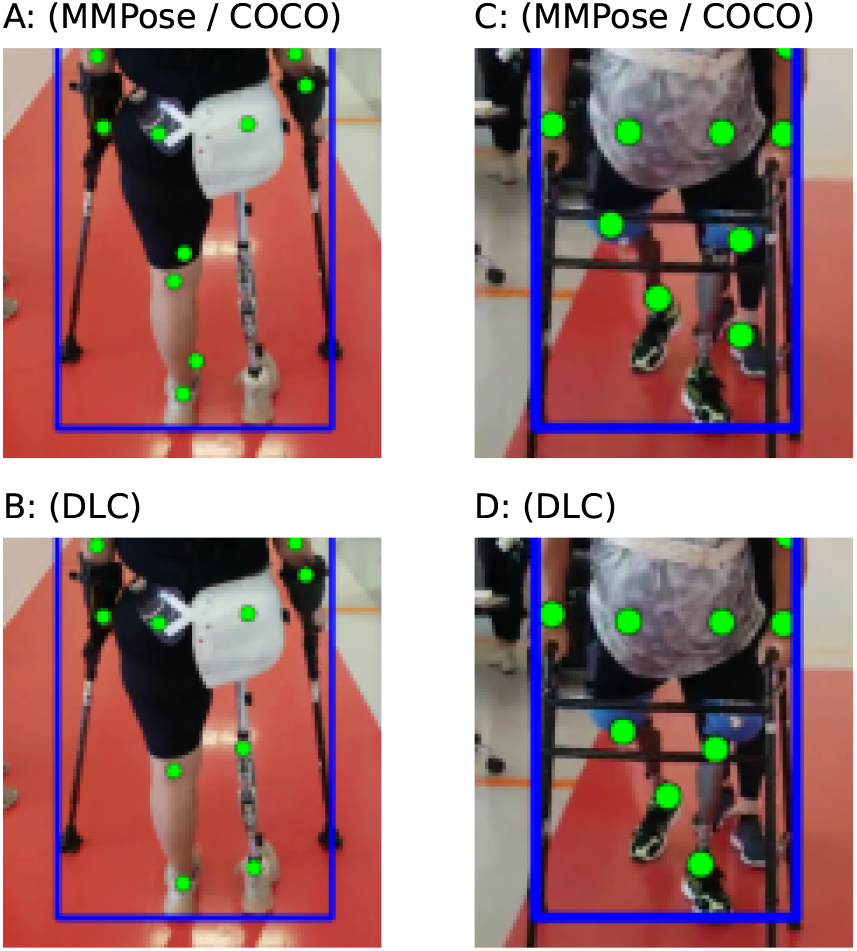
Examples of poor keypoint detection using pretrained algorithm (top row) that were corrected by training with DLC (bottom row)

#### Viewpoint sensitivity and clipping

We also classified segments as clipped or not if any of the 2D keypoints of the leg hit the edge of view on more than 1% of the frames, because we found the transformer was sensitive to the errors this produced. This occurred in none of the 79 frontal views, in 16 of the 67 sagittal views, and in 7 of the 85 mixed or oblique views. This was due to the somewhat limited space in clinical settings, where in hallways it can be hard to track far enough to the side of a person of interest to frame them with room for error.

#### Velocity and cadence accuracy

We compared the velocity estimated from the video acquired in the frontal plane with the gait transformer to the velocity computed from the manually annotated times as participants walked over a 10-meter interval between the tape on the ground. We found the gait transformer velocity for videos acquired in the frontal plane was quite accurate compared to ground truth, with a correlation (r) of 0.95 and a mean absolute error (MAE) of 0.14 m/s (**Fig 6a**). We also found that the cadence from the gait transformer was a close fit to the sensor data with an MAE of 4.8 steps/min, with most of this error coming from outliers (**Fig 6e**).

**Fig. 6.**
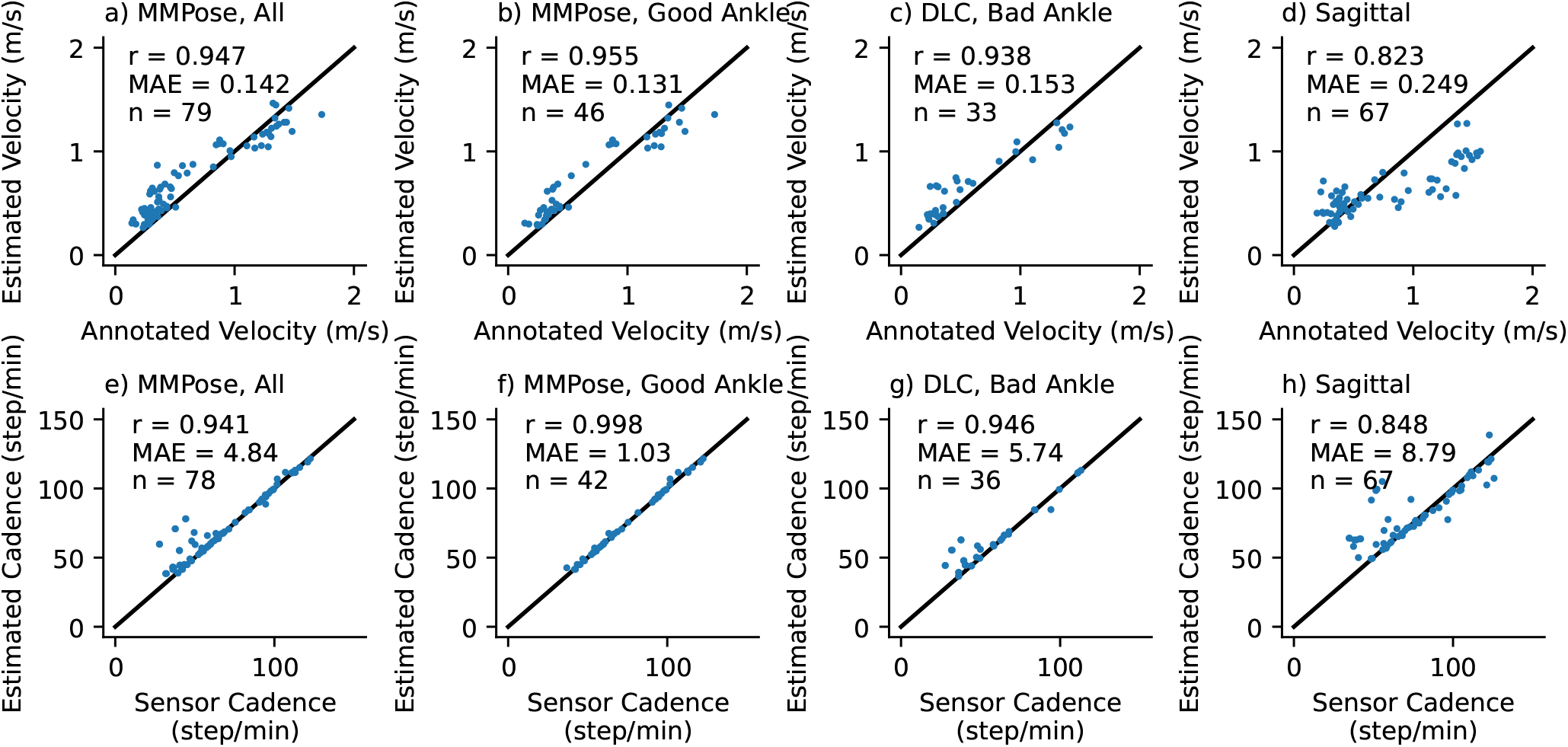
Accuracy of gait transformer for velocity (top row) and cadence (bottom row) under different conditions (columns). Text insets note the correlation coefficient (r), mean absolute error (MAE), and number of walking segments (n). The first column is all videos in the frontal plane. The second column is those where the average ankle detection quality was greater than 0.7. The third column is the excluded segments reprocessed with DLC. The last are videos acquired in the sagittal plane.

We repeated this analysis, excluding segments where the average prosthetic ankle quality was less than 0.7 (**Fig 6b,f**). Notably, we found this removed almost all the error estimating cadence (r=0.998, MAE=1.0). The accuracy of the estimated velocity also improved slightly (MAE=0.13 m/s). We then used our custom prosthetic keypoint detector trained in DLC to replace the prosthetic joints in the excluded segments (**Fig 6c,g**). We found this improved cadence detection for most sessions, but in particular, there were still several outliers for one individual with a slower cadence compared to other individuals.

We found the performance was quite poor when tested on video acquired in the sagittal plane (**Fig 6d,h**). This was relatively unsurprising since the gait transformer is trained on video acquired in the frontal plane. We attempted to improve the viewpoint invariance by augmenting the training process by randomly rotating the lifted 3D keypoints. We found that while it resulted in a tighter correlation with the outputs, both in the gait laboratory validation dataset and on the prosthetic data it resulted in a bias that underestimated velocity at faster speeds.

#### Step time accuracy

Over the frontal view timed walking segments, we detected and matched 889 prosthetic foot contact events and were unable to detect only 12 of these events. For each walking segment, we also computed the mean absolute error of the residuals. The average of this over all sessions was 72ms. For sessions with good ankle tracking, we detected 490 foot contact events and missed only 1 event, with an average error over sessions of 45ms.

## Discussion

Previously, we trained an algorithm for video-based gait analysis on a large clinical gait laboratory dataset of paired videos and motion capture data (24). While the algorithm validated well on this dataset, there are differences between the training data and the intended application of the algorithm that could result in poor generalization. These include recording portrait videos while walking with the patient through a therapy clinic where other people are present. Our training dataset also had a predominance of children, who are most commonly analyzed in these laboratories. For any clinical application of AI, it is critical to evaluate the external validity (or out-of-domain generalization in machine learning parlance) for the intended use case.

In this study, we evaluated the performance of our gait transformer when tested on prosthetic users walking in therapy or outpatient clinic. This was a powerful stress test for our algorithm, as there are several properties of prosthetic users that might cause a failure to generalize including the visual appearance of prostheses and prosthetic gait patterns. This study also highlighted the power of an interpretable pipeline, with meaningful features at multiple stages, which enables identifying and alleviating points of failure, such as 2D key-point detection accuracy or failures to detect a single step.

Large datasets are a critical driving force of AI algorithms and have enabled impressive advances in HPE in recent years. However, public HPE datasets, like COCO (33), primarily contain able-bodied individuals. Perhaps unsurprisingly, we found that a 2D keypoint detector trained on COCO did a poor job detecting the ankles of some prosthetic users. This was most pronounced with more proximal or bilateral amputations. This is most likely due the greater visual difference between these limbs and joints compared to the able-bodied individuals in the dataset, resulting in worse out-of-domain generalization of the algorithms. In these cases, tools like DLC (36) make it relatively easy to train a custom key-point detector. Importantly, this work also speaks to the need for more work on AI fairness for people with disabilities (37, 38).

When the 2D keypoints were accurately localized, we found that our algorithm performed well on videos of prosthetic users acquired in the frontal plane. Specifically, we had a mean absolute error for the velocity of 0.13 m/s and for the cadence of 1 step/min. Whether this is sufficiently accurate ultimately depends on the clinical question. One study found the minimally clinically important difference for walking velocity of prosthetic users as 0.21 m/s (39), which is greater than our algorithm. However, for older adults, it has been suggested a small meaningful change in walking speed is 0.05 m/s (40). Our results also do not indicate whether analyzing longer segments of walking would reduce the error, or whether this arises from a bias for given individuals that would persist over longer recordings.

The requirement to record video in the frontal plane to obtain accurate results is a limitation of our current approach. However, given the difficulties obtaining good videos in sagittal planes while walking in hallways and therapy gyms with other people present, it is also the most convenient approach for its intended use setting. An argument for sagittal videos is that they should enable more accurate estimates of many important sagittal plane kinematics. Our system outputs many of these and performs well on the training data but testing the external validity of these outputs is important future work. Specific to prosthetic users, testing the external validity of joint kinematics will be an area of challenge as prosthetic components attempt to mimic anatomic motion but do not move in an exact manner as able-bodied joints. As the gait transformer was primarily trained on individuals with intact limbs, this may affect the ability to accurately estimate prosthetic joint kinematics.

Several other groups have looked at the ability to estimate gait parameters from monocular video. Stenum and colleagues analyzed sagittal videos from a dataset (41) of 32 healthy individuals walking down a walkway a fixed distance from a mounted camera using OpenPose, and showed they could accurately estimate gait event timing, sagittal plane joint angles, step length and walking velocity (20). A pipeline similar to ours using 2D keypoints followed by lifted 3D joints was used with height-informed skeletal refinement prior to extracting gait parameters of healthy individuals walked towards a stationary camera and produced accurate estimates of step timing, length, and walking velocity (21). Similarly to our work, both of these computed features on individual gait cycles, but in comparison, they were not externally validated on clinical populations or on video acquired in the community or clinic. A neural network can also be trained to directly map 2D keypoint trajectories from the video onto average gait parameters, and such an approach has been used on gait laboratory data from children (22) and subsequently we showed a similar approach with stroke survivors (23). However, this approach does not allow for examining the cycle-by-cycle variability of gait parameters. Finally, by triangulating keypoints detected from multiple cameras it is possible to estimate more accurate 3D keypoint locations and perform inverse kinematics to fit biomechanical models to them (16, 17). OpenCap is a particularly portable version of this that only requires two iPhones (19).

There are several future directions that we anticipate will improve this system. One is better fusion with additional modalities of data, including the camera depth channel and inertial measurements from the wearable sensors. In this work, we have focused purely on monocular video as this is the most widely available modality. We are also enthusiastic about integrating physics-based modeling in the inference process (42–45) and ways to combine this with self-supervised learning (46). It is also important to make this system easier to use in a higher-throughput manner. One need is to automate the annotation step to robustly identify the subject of interest, such as through the use of QR codes. In this work, we used the periods where 10-meter walk tests were annotated to select what to analyze, but this is only a small fraction of the data we acquired. Utilizing activity recognition to identify when the subject is walking, using the detected pose to identify viewpoint, and the Kalman error to determine when walking is being reliably tracked could help automate analysis of the remainder of the data. We recently developed a 3D lifting algorithm that produces calibrated confidence estimates of the joint locations, and integrating this could also help determine the trustworthiness of outputs (47). Finally, there are many other clinically meaningful gait parameters available in our dataset that we could train the gait transformer to output and validate, including step width and center of mass.

## Data Availability

The primary data used in this work includes videos of patients including faces, and is not available.

## ACKNOWLEDGEMENTS

This work was generously supported by the Research Accelerator Program of the Shirley Ryan AbilityLab.

